# Discovery of a radiation countermeasure therapeutic for intestinal injury enabled by human organ chips combined with AI

**DOI:** 10.1101/2025.10.03.25337298

**Authors:** Alican Özkan, Gwenn Merry, Joshua Piatok, Arash Naziripour, Nina LoGrande, Thomas Matthiessen, Ryan R. Posey, Megan Sperry, Russel Gould, Kevin Ho, Agathe Neukelmance, Ela Contreas-Panta, Rocco Riccardi, Liliana Bordeianou, David Chou, David Breault, Girija Goyal, Donald E. Ingber

## Abstract

There is a need for better therapies for acute radiation injury (ARI) of the human intestine as current treatments offer limited efficacy. As the ileum is most sensitive to radiation in patients receiving cancer radiation therapy, we created human Organ Chip microfluidic culture models lined by primary patient-derived ileal epithelial cells interfaced with intestinal microvascular endothelium and exposed them to clinically relevant doses of γ-radiation. These Ileum Chips recapitulated key features of ARI, including cell loss, barrier dysfunction, and inflammation, as well as a therapeutic response to a probiotic formulation (VSL#3) that protects against radiation injury in patients. Use of an AI-enabled drug repurposing algorithm (NemoCAD) with transcriptomic data led to the identification of the antifungal agent miconazole as a potential radiation countermeasure drug, and its protective activity was confirmed on-chip. Combination of AI and human Organ Chip studies may offer a powerful way to repurpose drugs for novel disease applications.

**Highlights:**

- Primary human Organ Chips lined by patient-derived ileal epithelial cells interfaced with intestinal microvascular endothelium faithfully recapitulate acute radiation-induced intestinal injury
- Use of the Human Ileum Chip in combination with an AI-based drug repurposing platform led to the identification that the FDA approved antifungal drug miconazole has the potential to be rapidly repurposed as a therapeutic countermeasure against acute radiation injury in the human intestine.

## Introduction

Acute Radiation injury (ARI) of the intestine can occur after exposure to high doses of ionizing radiation can be a toxic side effect of radiation therapy that is administered to more than half of cancer patients(*1*) or result from an accidental or deliberate nuclear incident(*2*). While there are a few medical countermeasure (MCM) drugs approved for use in the treatment of ARI, they all target effects on the hematopoietic system. There is currently no therapeutic available for prevention of ARI of the intestine.

Radiation exposure causes DNA damage and stimulates free radical production in intestinal cells, which triggers an inflammatory response leading to epithelial cell damage, disruption of the crypt–villus architecture, mucus delamination and severe injury to the neighboring endothelium, particularly in rapidly proliferating regions of the intestine, such as the small intestine(*3*). The ileum is more susceptible to g-irradiation than the duodenum or jejunum, possibly due to relatively higher density of crypt stem cells and bacterial density as well as its location in the pelvic area (with respect to radiation therapy). This injury, which is called radiation-induced enteritis or enteropathy, occurs ∼1 week of exposure and is commonly diagnosed based on clinical symptoms, including abdominal cramps, diarrhea, rectal bleeding, and emesis(*4*). At present, most patients are given prolonged palliative therapy with corticosteroids (e.g., hydrocortisone, prednisolone, budesonide) and dietary modification (low fiber and residue diet) to reduce inflammation and small bowel irritation for 2-8 weeks after radiation exposure(*5, 6*). Unfortunately, there is limited evidence to support its effectiveness at reducing enteritis, although there may be some benefit in terms of preventing goblet cell depletion^7,8^. Therefore, there is a great need for the development of MCMs that specifically mitigate the effects of ARI in the intestine. However, pursuit of this goal has been held back by the lack of clinically relevant preclinical models.

Animal models have been employed to understand the effects of radiation on intestinal tissues, but their failure to mimic clinically relevant dose sensitivities and recapitulate key hallmarks of the human pathophysiology of ARI limit their value(*5*). Currently, non-human primate and mouse models are considered the gold-standard for ARI modeling, but their use is limited by short supply, long breeding periods, high costs, and serious ethical concerns(*7*). Human intestinal epithelial organoid cultures exposed to radiation have provided some insight to the effects of radiation on individual cell types(*8–11*), but they do not provide more physiologically relevant information about inflammatory responses, destruction of tissue barriers, and the interplay between different cell types that are observed in the human intestine at the organ level which are central to the development of radiation-induced enteritis(*12–14*).

In a past study, we modeled ARI in the intestine using organ-on-a-chip (Organ Chip) microfluidic culture devices(*15, 16*) lined with an established intestinal epithelial cell line originally isolated from a tumor (Caco-2 cells) interfaced with intestinal microvascular endothelium(*17*). To overcome the limitation of using an established cell line, here we used primary human Intestine Chips lined by primary ileal epithelial cells established from patient-derived ileal organoids interfaced with primary small intestinal microvascular endothelium, which experience dynamic fluid flow and cyclic mechanical deformations to mimic peristalsis-like intestinal motions, and we cultured these chips in the presence or absence of a complex living gut microbiome. We used this more physiologically relevant primary human Ileum Chip to both model ARI in the most sensitive region of the small intestine and identify potential MCM drugs that could mitigate these effects.

Our studies confirm that the primary human Ileum Chip can recapitulate many of the physiological hallmarks of intestinal radiation injury exposed to clinically relevant doses of γ-radiation and replicate the protective effects of a live biotherapeutic product that were previously demonstrated in human patients. More importantly, analysis of transcriptomics data obtained from these studies using the artificial intelligence (AI)-enabled drug repurposing algorithm NemoCAD (*18–20*) predicted that the FDA-approved antifungal drug miconazole should exhibit radioprotective effects in intestine. Indeed, when we tested miconazole in the human Intestine Chips, we confirmed that it significantly mitigated the damaging effects of ARI, and its protective effects appeared to be mediated by changes in matrix metalloproteinase (MMP)-9. To our knowledge, this represents the first radiation MCM drug that has demonstrated protective activity in human intestine.

## Results

### Primary human Ileum Chips recapitulate hallmarks of acute radiation injury

To build the primary human Ileum Chips, patient-specific epithelial organoids were isolated from histologically healthy regions of surgical resections or biopsies obtained from ileal regions of the intestines of cancer patients and cultured within the top channel of a 2-channel commercially available Organ Chip device on the upper surface of a porous extracellular matrix (ECM)-coated membrane that separates the top channel from a parallel lower channel and culture medium was perfused through both channels (**Fig. 1A**). After 14 days, when the epithelium was observed to form undulating villus-like structures, the intestinal microvascular endothelial cells obtained from a commercial supplier were seeded on the lower surface of the same membrane in the channel below (**Fig. 1A**) and flow was restored. Rhythmic mechanical deformations also were applied to the tissue-tissue interface to mimic peristalsis-like motions by applying cyclic vacuum to side chambers in the flexible device.

**Figure 1.**
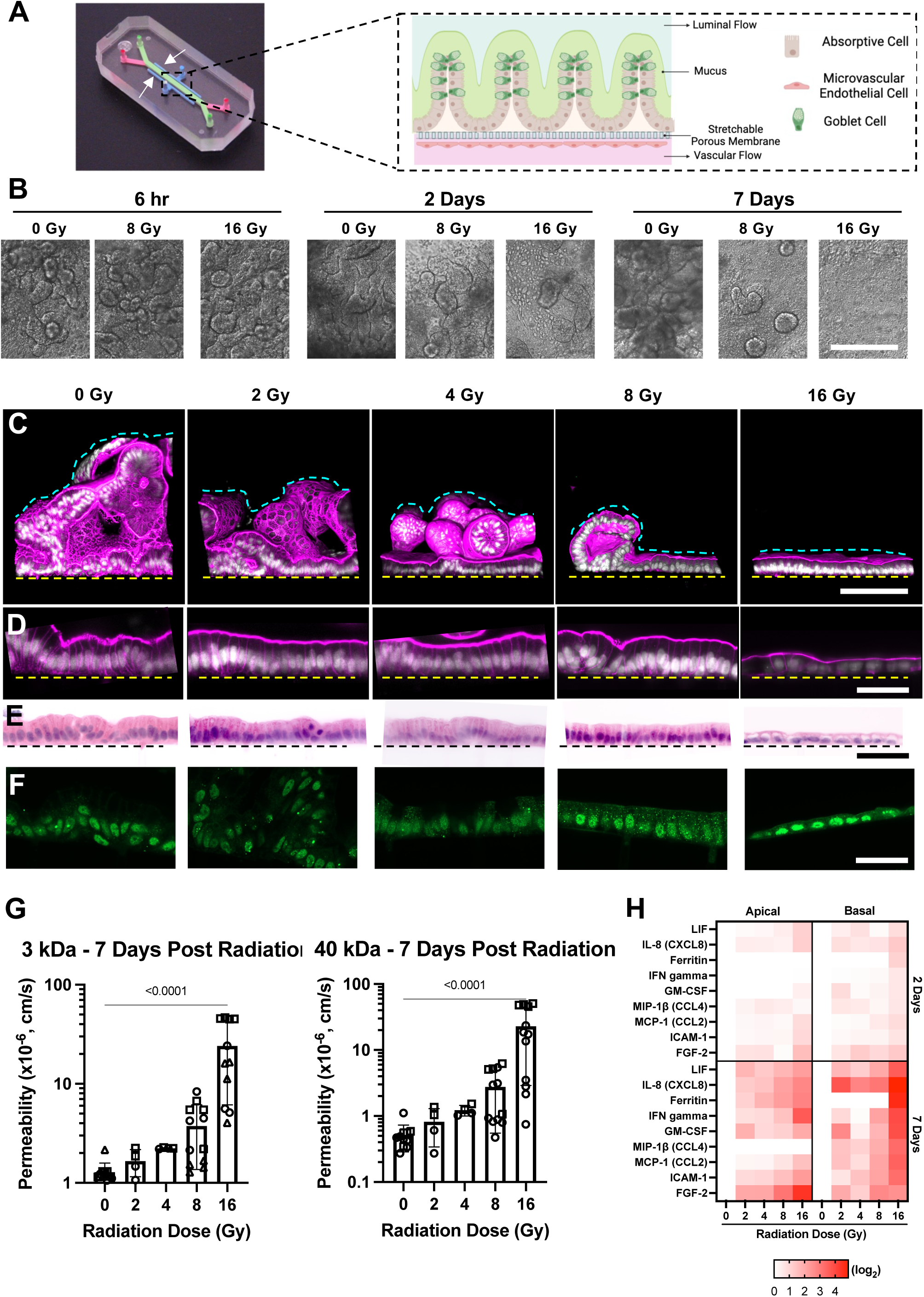
Human Ileum Chip recapitulates hallmark features of ARI. **A)** Photograph of an Organ Chip and a diagram showing how intestinal epithelial cells isolated from patient-derived ileal organoids and intestinal microvascular endothelial cells were cultured on opposite sides of a porous ECM-coated membrane that separates two parallel channels of a commercially available Organ Chip device to form epithelial-endothelium tissue interface. Culture medium is flowed through the upper epithelium-lined channel and lower endothelial channel to mimic movement of intestinal contents and vascular fluid flow, respectively. The epithelium,which is composed of absorptive cells and Goblet cells, forms villi-like structures and secretes a mucus layer into the lumen of the apical channel. The entire tissue-tissue interface can be stretched and relaxed rhythmically to mimic peristalsis-like motions by applying cyclic suction to hollow side chambers in the flexible device (shown with white arrows at left). **B)** DIC images of the villus epithelium when viewed from above in Ileum Chips after being exposed to radiation for 6 hours to 7 days. Radiation exposure results in a reduction in the number of villus-like structures (bar, 500 μm)**. C)** Immunofluorescence micrographs showing vertical cross-sections of Ileum Chips that demonstrate changes in the morphology of villus-like epithelial structures 7 days after different irradiation doses. Magenta, phalloidin-stained F-actin; white, DAPI-stained nuclei; Dashed yellow line, upper surface of chip membrane; dashed cyan line, upper luminal boundary of the villus (bar, 100 µm)**. D)** Higher magnification views of immunofluorescence micrographs from **C** showing regions of the ileal epithelium where it contacts the porous ECM-coated membrane on-chip (bar, 50 µm) **E)** Histological H&E stained vertical cross-sections confirming 16Gy radiation dose significantly reduces the height of the epithelial monolayer (bar, 50 µm). **F)** Immunofluorescence micrographs of vertical cross sections through the Ileum Chip showing 53bp1 staining (green) for double-stranded DNA breaks 7 days after exposures to different levels of radiation (bar, 50 µm). **G)** Graphs showing that intestinal barrier permeability measured with either 3 or 40 kDa Cascade Blue is significantly higher in Ileum Chips exposed to 16 Gy radiation for 7 days (n = 3 donors). Each symbol represents an individual health patient derived chip. **H)** Production of various pro-inflammatory cytokines and chemokines by Ileum Chips exposed to different radiation doses (n = 3 donors).

When these Ileum Chips were exposed to increasing doses of radiation from 0 to 16 Gray (Gy), we found that the number of villus-like epithelial extrusions and their height decreased in a dose-dependent manner at 2 and 7 days after irradiation when viewed from above using differential interference microscopy (DIC) or from the side in vertical cross sections by immunofluorescence microscopy, with the epithelium appearing fully blunted by day 7 when exposed to the highest 16 Gy dose (**Fig. 1B,C**). Computerized morphometric analysis confirmed that there was a radiation dose-dependent reduction in villus height on days 2 and 7 (**Extended Data Fig. 1A**). We also observed a change in epithelial cell morphology as the normally columnar epithelium flattened in response to exposure of increasing radiation doses (**Fig. 1D,E**). This was accompanied by a similar dose-dependent increase in DNA damage in the ileal epithelium as measured by quantifying punctate 53bp1 staining in nuclei (**Fig. 1F**), which correlates with formation of DNA double strand breaks (DSBs)(*21*). Using small (3kDa) and large (40 kDa) fluorescent tracers, we found that the intestinal tissue barrier remained intact until 2 days after radiation exposure, but barrier compromise with permeability increasing significantly (>10-fold) was detected at the16 Gy exposure on days 4 to 7 (**Fig. 1G**, **Extended Data Fig. 1B**). Intestinal inflammation is also a common feature of ARI in humans, and we also found more proinflammatory cytokines and other inflammatory mediators, including LIF, IL-8, ferritin, IFN-γ, GM-CSF, MIP-1β, MCP1, ICAM-1, and FGF-2, in effluents of both the epithelial-lined (apical) and endothelial-lined (basal) channels of the device when exposed to progressively higher radiation doses (**Fig. 1H**). Again, the greatest increases in these inflammatory modulators were observed at 7 days, which correlates with when human patients experience radiation-induced enteritis.^20^ Another hallmark of ARI to the intestines is decreased mucus production in vivo(*22*) and we observed a similar dose-dependent reduction in the accumulation of dense opaque deposits covering the intestinal epithelium on-chip by day 7 (**Extended Data. Fig.1C**). We have previously shown that this dense opaque material that accumulates in the upper channel on-chip is mucus with similar composition, thickness, and bilayer structure as seen in human small intestine in vivo(*23–25*).

Consistent with the greater effects on intestinal barrier, inflammation, and mucus being observed at 7 days, we also found that there were minimal changes in gene expression (< 40 genes) in either the epithelium or endothelium at 6 hour post radiation compared to controls, while more than 1,000 genes were differentially expressed at 1 week after exposure to 16 Gy (**Fig. 2A**). Not surprisingly, pathway analysis revealed that biological processes associated with cytokine production, cell death, and p53 signaling pathway were activated while cell proliferation-associated pathways were suppressed (**Fig. 2B**). Mutual information analysis revealed that multiple radiation-associated radiation injury-associated genes (e.g. FDXR, MDM2, CDKN1A, BAX, BACE2, H3C4) were enriched in both epithelial and endothelial cells cultured in irradiated Ileum Chips (**Fig. 2C**).

**Figure 2.**
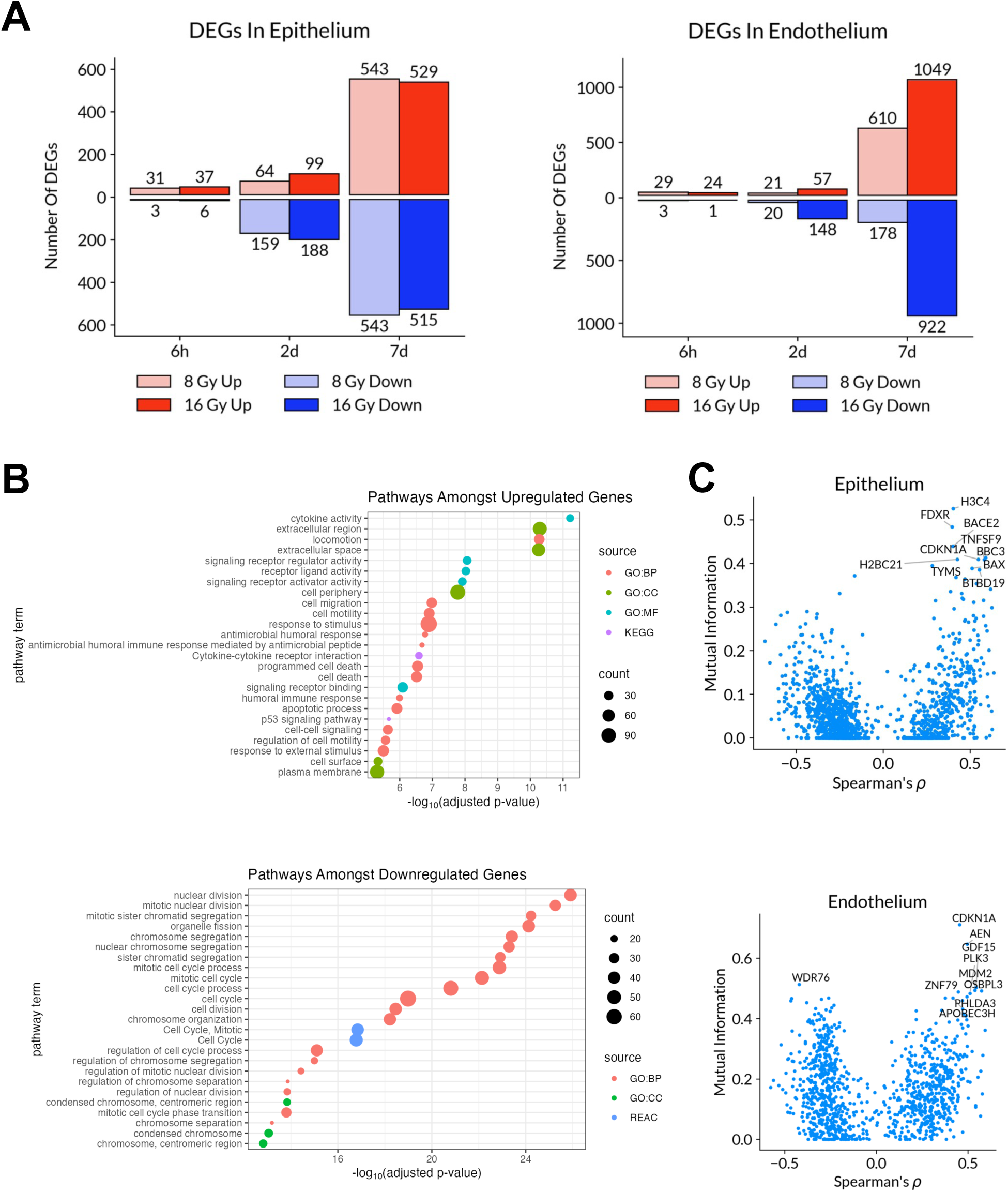
Transcriptomic analysis of the effects of radiation exposure in human Ileum Chips. **A)** Numbers of significantly up- and downregulated DEGs in Ileum Chips in response to 8 or 16 Gy irradiation at 6 hours, 2 days, and 7 days post-irradiation. The number of DEGs increased with time, with far fewer DEGs at 6 hours and 2 days compared to 7 days. **B)** Analysis of molecular pathways affected by the most significantly upregulated and downregulated genes at 6 hours, 2 days, and 7 days post-irradiation with 8 Gy and 16 Gy in human Ileum Chips. **C)** Top differentially expressed genes in epithelial and endothelial compartments of irradiated Ileum Chips, highlighting expression of DSB markers and radiation biomarkers (n = 3 donors).

### Human Ileum Chips recapitulate the clinical response to a live biotherapeutic product

The gut microbiome has been shown to suppress enteritis and endothelial apoptosis in response to exposure to lethal radiation in mice. More importantly, a commercially available live biotherapeutic product composed of a probiotic consortium composed of multiple bacteria found in the intestine of healthy patients known as VSL#3 has been reported to reduce radiation-induced gastrointestinal symptoms in human patients (*26–28*). As we have previously shown that complex, living, human gut microbiome can be cultured in the epithelial lumen of human Intestine Chips under low oxygen conditions for days without interrupting tissue barrier integrity(*29*), we explored whether the Ileum Chip could model this clinical response to this radiation countermeasure drug. The apical lumen of the Ileum Chips was inoculated with complex microbiome isolated from stool obtained from healthy newborn human infants in the absence or presence of the VSL#3 probiotic consortium 24 hour prior to the chips being exposed to 8 Gy radiation. Control studies showed that culture with VSL#3 alone was well tolerated by the Ileum Chips for at least 3 days, as indicated by the maintenance of tissue barrier integrity (**Extended Data Fig. 2A**) and absence of production of proinflammatory cytokines (**Extended Data Fig 2B**). In contrast, the presence of the complex human gut microbiome alone was found to exacerbate the radiation-induced decrease in villus height when viewed from above by DIC optics (**Fig. 3A**), in histological cross-sections (**Fig. 3B**), or when height was quantified (**Fig. 3C**). In contrast, this was not observed when chips containing VSL#3 alone were irradiated, and more importantly, the presence VSL#3 mitigated the additional villus blunting effect induced by the presence of a complex gut microbiome (**Fig. 3A-C**).

**Figure 3:**
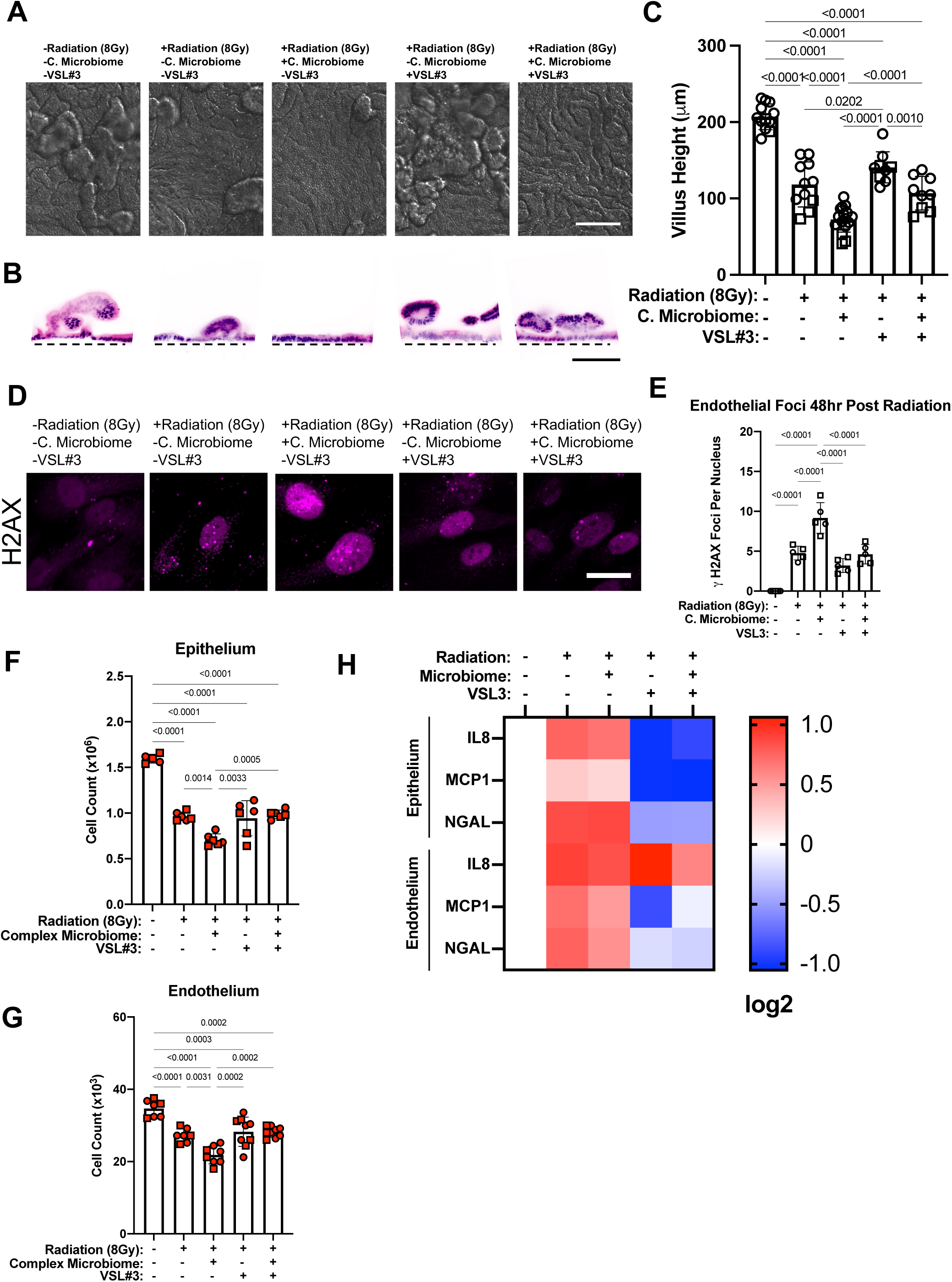
VSL#3 prevents tissue-resident intestinal complex microbiome’s contribution to radiation-induced epithelium and endothelium damage in primary human Ileum Chip. **A)** DIC images of the epithelium viewed from above in Ileum chips, 2 days post radiation in the presence or absence of complex microbiome or probiotic VSL#3 consortium (Bar, 100 µm). **B)** Histological H&E stained vertical cross-sections of Ileum Chips showing that the complex microbiome can exacerbate radiation-induced villus blunting (Bar, 100 µm). Dashed black line, upper surface of chip membrane. **C)** Quantification of results from **B** demonstrates that VSL#3 prevents the contribution of complex microbiome to radiation-induced villus blunting in Ileum Chips at 2 days post radiation. **D)** Confocal images of endothelial cells cultured in Ileum Chips stained for H2AX 48 hours post radiation exposure with complex microbiome, VSL#3 or both (Bar is 10 µm) **E)** Quantified results from **D** confirmed that VSL#3 can prevent complex microbiome’s exacerbation in radiation-induced nuclear H2AX foci number increase in endothelial cells cultured in Ileum Chips. **F)** Quantified epithelial cell number reduction 48 hours after chips were exposed to radiation when cultured with complex microbiome, VSL#3 or both. **G)** Decrease of endothelial cell number in Ileum Chips induced by complex microbiome can be prevented with VSL#3 probiotic consortium 48 hours post radiation. **H)** VSL3# can partially prevent radiation-induced inflammatory cytokine and chemokine production in apical and basal channel outflows. Presence of VSL#3 prevents radiation-induced IL-8, MCP-1 and NGAL secretion into the apical channel. VSL#3 prevents radiation-induced MCP-1 and NGAL secretion but not IL-8 into the basal channel. Data represents 2 donors from 3 independent experiments. Numbers indicate P values between compared groups, as determined by one-way ANOVA test (n=2 Healthy donors indicated by different symbols from 3 independent experiments. Each symbol represents an individual health patient derived chip.

Analysis of the intestinal microvascular endothelium in these chips revealed that it exhibited greater DNA damage as indicated by staining for nuclear H2AX when irradiated in the presence of the complex microbiome, and this too was prevented when VSL#3 was included in the model (**Fig. 3D,E**). Time-course analysis of DNA double stranded breaks in endothelial cells cultured in these chips revealed that the expression of both H2AX and 53BP1 increased 1 hour after radiation and gradually decreased over the following 24 hours (**Extended Data Fig. 3A, B**). Interestingly, culturing VSL#3 in Ileum Chips before irradiation did not prevent the decrease in epithelial and endothelial cell number caused by exposure to 8 Gy radiation for 2 days; however, it prevented the additional reduction in cell number that resulted when a complex gut microbiome was present (**Fig. 3F,G**). Cytokine analysis also showed that the presence of VSL#3 can reduce radiation-induced NGAL and MCP-1 production in both apical and basal channel effluents even though this effect was not augmented by the presence of the complex microbiome; however, VSL#3 only reduced IL-8 production in the epithelial channel (**Fig. 3H**).

### AI-enabled discovery of a novel radiation countermeasure drug

Having generated transcriptomic datasets from human intestinal epithelium and endothelium within Ileum Chips cultured in the presence or absence of exposure to a 16 Gy dose of γ-radiation that replicated many of the hallmarks of radiation-induced enteritis observed in humans over a similar time course, we set out to explore if we could use a recently developed artificial intelligence (AI)-based drug repurposing algorithm to identify novel radiation countermeasure drugs specifically for ARI in the human intestine. These transcriptomic data were analyzed using the Network Model for Causality-Aware Discovery (NeMoCAD) computational tool that combines AI (machine learning) and gene network analysis to predict drugs that would shift the ARI transcriptomic state to a healthy state using interaction probabilities of drug-gene and gene-gene interactions based on analysis of differential gene expression signatures of the disease state versus healthy state. NeMoCAD has been previously used successfully to identify drugs that may be repurposed for other medical conditions, ranging from COVID-19 to Rett Syndrome (*18–20*). This analysis resulted in production of a list of compounds that are predicted to be potential countermeasures against irradiation, which included the antifungal drug miconazole.

To explore whether miconazole has radiation protective effects, we administered miconazole to Ileum Chips through their apical or basal channel to recapitulate oral and intravenous delivery, respectively, 30 minutes after exposure to 16 Gy irradiation. Indeed, treatment with both 0.1 and 1 µM miconazole partially prevented the decrease in intestinal villi (**Fig. 4A**) as well as blunting of the villus and epithelial cell heights (**Fig. 4B,C,D**) and the compromise of the tissue permeability barrier (**Fig. 4E**), regardless of the delivery channel or the concentration used. Miconazole treatment also prevented the radiation-induced reduction in intestinal mucus (**Fig. 4F,G**) and increase in production of inflammatory cytokines (**Fig. 4H**); however, perfusion of the drug through the apical (epithelial) channel was more effective at producing these effects. Interestingly, treatment with miconazole in the basal channel also resulted in the upregulation of several pro-inflammatory cytokines, including IL-8 and NGAL in both channels, and IL-10, IFN-γ, and MIP-1β specifically in the vascular channel. Treatment of irradiated Ileum Chips with miconazole also reduced radiation-induced VE-cadherin junction disruption (**Extended Data Fig. 4A**), and number of 53bp1 localized in nucleus of endothelial cells (**Extended Data Fig. 4B, C)**

**Figure 4:**
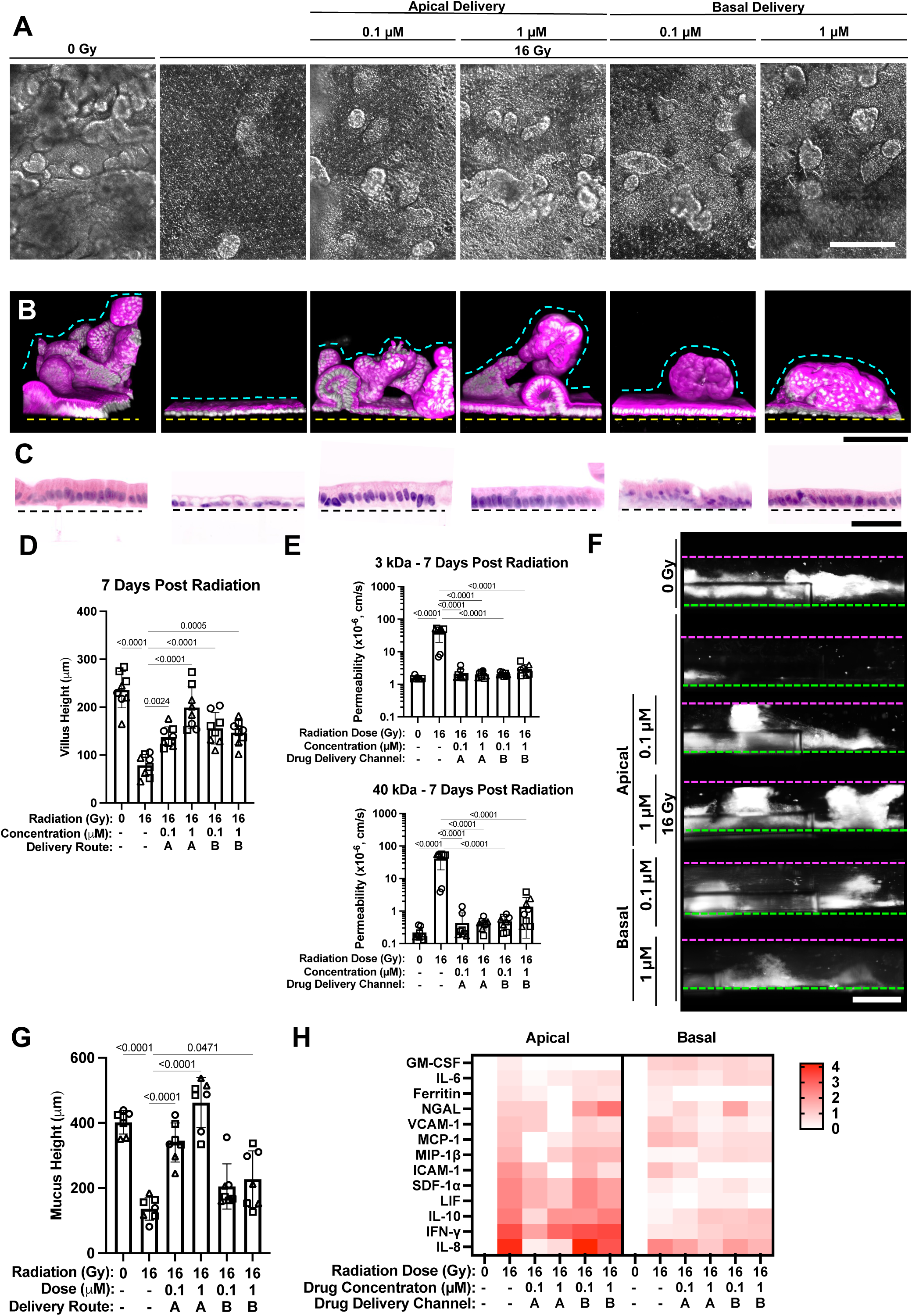
Miconazole Treatment Prevents ARI-Induced Associated Phenotypes in Ileum Chips. **A)** DIC images of the epithelium when viewed from above in Ileum Chips 7 days after being exposed to 16 Gy irradiation and treated with miconazole for 7 days. **B)** Immunofluoresence vertical cross-section micrographs of Ileum Chips showing the villus-like epithelial structures 7 days after 16 Gy irradiation and treatment with miconazole. Magenta, phalloidin; white: DAPI; Dashed yellow line, upper surface of chip membrane; dashed cyan line, upper luminal boundary of the villus (bar, 100 µm)**. C)** Histological H&E stained vertical cross-sections confirming miconazole treatment prevents 16Gy radiation dose-induced reduction of epithelial monolayer height (Bar, 50 µm). **D)** Quantification of villus height from confocal images shows that miconazole treatment reduces radiation-induced villus blunting in a dose- and delivery route dependent manner. **E)** Quantification of barrier permeability to 3 kDa and 40 kDa Cascade Blue tracers demonstrates that miconazole mitigates radiation-induced intestinal leakiness (n = 3 donors). **F)** Side-view images of Ileum Chips stained WGA reveal preservation of the mucus layer (white) 7 days after 16 Gy irradiation and apical miconazole treatment. Dashed green line, upper surface of chip membrane; dashed magenta line, upper boundary of the apical channel. **G)** Quantification of mucus height confirms that apical delivery of miconazole prevents radiation-induced mucus delamination. **H)** Cytokine and chemokine analysis of apical and basal outflows shows that miconazole treatment markedly reduces radiation-induced inflammatory responses after 7 days (n = 3 donors). Each symbol represents an individual health patient derived chip.

While miconazole is believed to primarily target CYP51 in cells, its off-target effects, including MMP9 inhibition and inflammation reduction, have been previously described in zebrafish and human cell lines (*30, 31*). Interestingly, we found that when we treated ileum organoids with a broad-spectrum MMP inhibitor (with ilomastat) or a highly selective MMP9 inhibitor (JNJ0966) following 16 Gy irradiation both improved cell survival to a similar degree as miconazole and combining these drugs did not have an additive effect (**Extended Data Fig. 4D**). This initial finding suggests that miconazole’s protective mechanism against ARI to the human intestine may be mediated at least in part by its ability to inhibit MMP9 activity.

## Discussion

ARI of the human intestine is a serious complication of high-dose radiation exposure and current treatments, including steroids, dietary modifications, and probiotic therapies offer limited efficacy. In this study, we leveraged microfluidic human Organ Chip technology to model ARI in the human ileum and combined it with an AI-based computational approach to identify an existing FDA approved drug - miconazole - that may be repurposed as a novel radiation countermeasure therapeutic. The human Intestine Chips recapitulates clinically relevant acute responses to radiation exposure at the cell, tissue, and organ levels, including cell loss, villus blunting, reduced mucus production, and compromise of intestinal tissue barrier integrity. Our data show that both the ileal epithelium and endothelium are sensitive to radiation injury in terms of pro-inflammatory cytokine and chemokine production and transcriptional changes. Moreover, pre-treatment with a known live biotherapeutic product, VSL#3, significantly reduced complex microbiome’s exacerbation of radiation toxicity in epithelium and endothelium in this in vitro model, again mimicking responses that have been previously observed in vivo, including in human clinical studies(*27, 32, 33*).

Importantly, we demonstrated that the Ileum Chip radiation model can be integrated with an AI-based drug repurposing algorithm that uses transcriptomic data as inputs so that potential radiation countermeasure drugs identified with AI can be evaluated for their therapeutic capacity in a human-relevant experimental system. As a proof-of-concept, we showed that transcriptomic data generated from irradiated and non-irradiated Ileum Chips combined with use of the NemoCAD drug repurposing AI algorithm predicted that the antifungal drug, miconazole, may protect against ARI in the intestine. Importantly, when we experimentally tested miconazole in the human Ileum Chip, it was found to demonstrate significant radiation countermeasure activity. To our knowledge, this is the first demonstration that AI and human Organ Chips can be integrated to both discover a new potential therapeutic and validate it efficacy.

In addition to demonstrating micronazole’s therapeutic efficacy, use of the Organ Chip approach also enabled us to some insight into its potential mechanism of action. Miconazole and other anti-fungals (e.g., fluconazole, itraconazole, voriconazole, amphotericin B, nystatin) are believed to target cytochromeP450-51 (CYP51). However, we found that miconazole’s ability to protect against ARI in the Ileum Chip was based largely on its ability to inhibit MMP9. Radiation exposure induces MMP9 mRNA expression in vivo(*34*) and administration of another compound that protects against intestinal radiation injury in a mouse model (rebamipide) similarly reduced MMP9 production(*35*). Miconazole treatment in our human Ileum Chips was most effective when delivered through the apical channel, which simulates oral administration, particularly in terms of mucus retention. This finding is particularly important because miconazole is currently available as oral tablets or topical formulations.

In summary, this work shows the power of combining AI with human Organ Chip technology in terms of both identifying therapeutics for new disease applications, and providing a human-relevant model that can be used to rapidly validate their efficacy experimentally. The discovery of the novel radiation protective activity of the antifungal miconazole demonstrates the proof-of-concept of this combined approach. One limitation of this study is that miconazole’s efficacy was not tested either alone or in combination with VSL#3 in Ileum Chips with complex microbiome; however, this could be tested using this method in the future. While we only demonstrated miconazole’s ability to protect against ARI in this human ileum model, it also should be explored in other organ systems. Given that we found that miconazole’s effects on MMP9 appear to be central to its radiation protection activity, it might be interesting to explore whether MMP9 also plays a role in its antifungal actions in the future.

## Methods

### Cell culture

Organoids from donors were generated from biopsy samples collected during exploratory gastroscopy (from Boston Children’s Hospital) or surgical resections (from Massachusetts General Hospital)(*36*). Informed consent and developmentally appropriate assent were obtained at Massachusetts General Hospital and Boston Children’s Hospital from the donor or their guardians. All methods were performed in accordance with the Institutional Review Board of Massachusetts General Hospital approval (protocol number IRB-2015P001859) and Boston Children’s Hospital (protocol number IRB-P00000529). We generated epithelium-derived ileum organoids that were cultured in Matrigel using published methods(*24, 37*). For isolation of human ileum cells from patient tissue specimens, tissues were processed by removing epithelium with lamina propria and digesting the entire specimen with 2 mg/mL collagenase I (17100-017;

Thermo Fisher Scientific, Waltham, MA) supplemented with 10 μmol/L Y-27632 (Y0503; Sigma-Aldrich, St. Louis, MO) for 30 min with occasional agitation. Organoids were grown embedded in growth factor reduced Matrigel (356231, lot 7317015; Corning, Corning, NY), and the intestinal expansion medium (EM) was composed of Advanced Dulbecco’s modified Eagle medium F12 (12634-010; Thermo Fisher Scientific) containing L-Wnt3a, R-spondin, noggin–conditioned medium (65% vol/vol) (produced by the CRL-3276 cell line; American Type Culture Collection, Manassas, VA), 1N GlutaMAX (35050-061; Thermo Fisher Scientific), 10 mmol/L HEPES (15630-106; Thermo Fisher Scientific), recombinant murine epidermal growth factor (50 ng/mL) (315-09; Peprotech, Rocky Hill, NJ), 1N N2 supplement (17502-048; Thermo Fisher Scientific), 1N B27 supplement (12587001; Thermo Fisher Scientific), 10 nmol/L human (Leu15)-gastrin I (G9145; Sigma-Aldrich), 1 mmol/L N-acetyl cysteine (A5099; Sigma-Aldrich), 10 mmol/L nicotinamide (N0636; Sigma-Aldrich), 10 mmol/L SB202190 (S7067; Sigma-Aldrich), 500 nmol/L A83-01 (2939; Tocris, Bristol, UK), Primocin (100 mg/ mL) (ant-pm-1; InvivoGen, San Diego, CA), which was supplemented with 10 μmol/L Y-27632 (Y0503, Millipore Sigma,MA).

Small intestine microvascular endothelial cells (HSIMECs, 10HU-065, iXCells Biotechnologies, CA) were expanded in endothelial basal medium-2 (Lonza CC-3156) supplemented with Primocin and bullet kit (Lonza CC-4176), which contains FBS, Hydrocortisone, hFGF-B, VEGF, R3-IGF-1, Ascorbic Acid, hEGF, Heparin. All cells were used up to passage 6.

### Ileum Chips

Our methods for creating and culturing human Ileum Chip lined by primary intestinal organoid-derived epithelium using commercially available Organ Chip Devices (Basic Research Kit; Emulate Inc.) have been described previously(*25, 38*). Optically clear Organ Chips made of poly-dimethylsiloxane (PDMS) and containing two parallel microchannels (top, 1 x 1 mm; bottom, 1 x 0.2 mm) separated by 50 μm thick PDMS porous membrane (7 μm pore diameter, 40 μm spacing) were purchased from Emulate Inc (Basic Research Kit). Surfaces were activated using 0.5 mg/mL sulfo-SANPAH solution (A35395; Thermo Fisher Scientific) for 20 min and coated with 0.2 mg/mL collagen type I (354236; Corning) and 1% Matrigel (356231, Corning) in Dulbecco’s phosphate-buffered saline (DPBS). During the coating step, organoids were obtained by dissolving Matrigel using cell recovery solution (Corning, 354253) for 40 min on ice and spun down at 400*g* for 5 min at 4° C. Organoids were then fragmented with TrypLE Express Enzyme (12605010; Thermo Fisher Scientific), diluted 1:1 in DPBS, and supplemented with 10 μmol/L Y-27632 (2 mL/well of a 24-well plate) for 2 min at 37° C. After adding the same amount of EM with 10 μmol/L Y-27632, Organoids were spun down at 400*g* for 5 min at 4° C and resuspended at 6 x 10^6^ cells/mL for seeding. Fragmented organoids were seeded in the apical channel of the 2-channel Organ Chip devices and incubated overnight at 37° C under 5% CO_2_ to promote adhesion. One day later, channels were washed with Expansion Medium (EM) to remove unattached cells, and then perfused with EM (60 µL/hr). After 3 days of culture during which epithelial monolayer was established, cells were exposed to physiological peristalsis-like motions, cyclic mechanical strain (10% Strain 0.15 Hz) by applying cyclic suction to side chambers of the flexible device(*29, 38*) for 2 weeks after seeding. After 2 weeks of epithelial cell culture, the basal channel was coated with 200 µg/ml collagen IV and 15 µg/ml laminin at 37 °C in 5% CO₂ for 2 hours. The bottom channel was then washed with PBS (−/−), followed by seeding with HSIMECs at a concentration of 8 × 10⁶ cells/mL. Two hours after seeding, the chips were reattached to the pods. For an additional 2 days, the apical medium was replaced with differentiation medium (DM composition), while the basal medium was maintained as epithelial medium (EM) supplemented with endothelial cell growth factors (Lonza CC-4176).

### Microbiome Preparation and Inoculation of Intestine-Chips

Stool-derived complex microbiome preparation and inoculation was prepared as previously described(*29*). VSL#3® probiotic capsules (Actial Farmaceutica Srl, Italy; 112.5 × 10⁹ CFU per capsule guaranteed at expiration; stored at 2–8 °C; purchased via Amazon, USA) were used as a countermeasure against radiation-induced ileal injury in Emulate Intestine-Chips seeded with human ileal organoid-derived epithelial cells. To recapitulate the anaerobic luminal microenvironment and maintain probiotic viability, all bacterial handling and inoculation steps were conducted within a hypoxic workstation (Coy Laboratory Products; 1% O₂, 5% CO₂, balance N₂). The procedure was adapted from established protocols for probiotic administration in gut-on-a-chip models of radiation injury (*39, 40*). Reinforced Clostridial Medium (RCM; Oxoid, cat. no. CM0149) and Lactobacilli MRS Broth (Difco, cat. no. 288130) were prepared according to the manufacturer’s instructions, autoclaved separately (121 °C, 15 min), and mixed 1:1 (v/v) under aseptic conditions (Class II biosafety cabinet). The mixed MRS:RCM broth (25 mL) was then transferred to the hypoxic workstation and pre-reduced overnight (12–18 h, 37 °C) with caps loosened to facilitate deoxygenation. On the following day, one capsule of VSL#3 was aseptically opened inside the workstation, and the powder was resuspended in 10 mL of pre-reduced MRS:RCM medium by gentle pipetting (10 cycles), followed by 5 min of settling to remove insoluble excipients. The suspension was incubated statically at 37 °C for 12–18 h to enrich and activate the probiotic consortium (*Streptococcus thermophilus*; *Bifidobacterium breve*; *B. longum*; *B. infantis*; *Lactobacillus acidophilus*; *L. plantarum*; *L. paracasei*; *L. delbrueckii* subsp. *bulgaricus*). That evening, Dulbecco’s Phosphate-Buffered Saline (DPBS; Gibco, cat. no. 14190-144) and Hank’s Balanced Salt Solution (HBSS; Gibco, cat. no. 14025-092) were also transferred into the hypoxic workstation and pre-reduced under identical conditions (12–18 h, 37°C). On the next day, 1 mL of the enriched bacterial culture was centrifuged at 4500 × g for 8 min, the supernatant discarded, and the pellet resuspended in 1 mL pre-reduced DPBS. This wash was repeated once, and the final pellet was resuspended in 1 mL pre-reduced DPBS. Serial tenfold dilutions were prepared in pre-reduced DPBS to at least 10⁻⁵ for plate enumeration. Based on the plate-derived CFU/mL of the enriched culture (typically 1×10⁹–5×10⁹ CFU/mL), a working inoculum dilution of ≈10⁻² (fine-tuned from the measured titer) was selected to yield ≈6.7×10⁷ CFU/mL. The working inoculum was prepared in pre-reduced HBSS; a 30 μL dose therefore delivered ≈2×10⁶ CFU per chip (MOI ≈ 2.5), consistent with prior studies (*39, 41*).

### Quality control of inoculum

Viable CFU/mL of the inoculum was determined by plating serial dilutions (100 μL aliquots) on pre-reduced MRS agar supplemented with 0.05% L-cysteine under anaerobiosis (37 °C, 48–72 h; plates held ≥24 h in the workstation before use). The expected viable count was 1×10⁹–5×10⁹ CFU/mL; if viability fell outside this range, the inoculum dilution was recalculated to achieve an MOI of ≈ 2.5 per chip before inoculation(*40*).

One day before inoculation, the apical (top) channel was switched to antibiotic-free HBSS, and the basal (bottom) channel was perfused with antibiotic-free complete expansion medium under standard flow (60 μL/h) and cyclic strain (10% amplitude, 0.15 Hz) to clear residual antibiotics. On the day of inoculation, antibiotic-free expansion medium was transferred into the hypoxic workstation and first perfused through the basal channel to maintain epithelial oxygenation.

Inside the workstation, 30 μL of the final bacterial suspension (in pre-reduced HBSS) was gently introduced into the apical channel to prevent disturbance of bacterial positioning during basal perfusion. Chips were then incubated statically under hypoxia (1% O₂, 37 °C) for 30 min to enable bacterial settling. Following this, chips were reconnected to the Pod reservoir (Emulate, Inc.), mounted on the Zoe™ Culture Module (housed within a normoxic incubator; 37 °C, 5% CO₂, atmospheric O₂), and held static for 1 hour to allow bacterial adherence. The Zoe was configured to maintain a hypoxic apical (luminal) channel and a normoxic basal (vascular) channel, after which physiomimetic flow and cyclic mechanical strain were resumed (60 μL/h; 10% amplitude, 0.15 Hz) for the remainder of the experiment (*39, 41*)

### Gamma Irradiation

Intestine Chips or cultured plates were placed in gamma irradiation chamber and radiated at 0.86 Gy/min rate at different duration of time to reach 2, 4, 8 or 16 Gy radiation doses. Control chips were placed outside of the incubator during this duration. Chips were then washed with warm HBSS in apical channel and EM in the basal channel to remove radiated media. For microbiome-related studies, 24 hours post inoculation of complex microbiome, VSL#3 or combined, Intestine Chips were irradiated at 8 Gy.

### Analysis of intestinal barrier integrity

To assess paracellular epithelial leakiness across the intestinal barrier in ileum chips, Cascade Blue (3 and 40 kilodaltons) (Invitrogen D7132 and D1829) were passed through the apical channel at 50 μg/mL. Initial and final outflows of both channels were collected, and luminescence was measured according to the manufacturer’s protocol. Permeability was calculated according to the following equation:

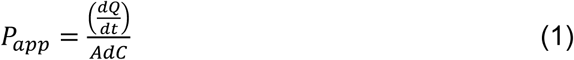

where A is the total area of diffusion, dC is the average concentration gradient and dQ/dt is molecular flux, as previously described(*25*)

### Morphological Analysis

Top-view images of the Intestine Chips were acquired using differential interference contrast (DIC) or phase-contrast microscopy (ECHO, San Diego, CA). Fixed samples that were sectioned (200 μm thick) with a vibratome (Leica, VT1000-S) were used for H&E staining to visualize epithelial structures.

For immunofluorescence imaging, Intestine Chips were fixed with 4% paraformaldehyde (50-980-487) in PBS(+/+) for 30 minutes at room temperature. Chips were either stained directly or sectioned at 200 µm thickness (Leica). All samples were blocked and permeabilized using 0.1% Triton X-100 (Sigma, X100-5ML) and 5% bovine serum albumin (BSA, A9418-50G) in DPBS for 1h at RT. Samples were incubated with primary antibodies in 2% BSA in DPBS overnight. The next day, secondary antibodies were incubated overnight. Finally, samples were counterstained with Hoechst (94403-1ML) for 30 min at RT. Samples were imaged using Zeiss TIRF/LSM 980 confocal microscope with lasers including 352, 488, 561, and 647 coupled with HyD detectors. Acquired images were analyzed either with ImageJ or IMARIS software (Biplane, Zurich, Switzerland). A list of primary and secondary antibodies used is provided in Table S1.

The height of extended epithelial villus-like structures was measured using confocal images of vibratome sectioned ileum chips. Villus-like structure height was quantified by measuring the maximum distance between the upper surface of chip membrane (visualized with dashed yellow line in the confocal images) and upper luminal boundary of the villus-like structures (dashed cyan line in the confocal images). ImageJ was used to quantify the distance between the membrane and epithelium.

Mucus of live chips was visualized using a wheat germ agglutinin (WGA)–Alexa Fluor 647 conjugate (ThermoFisher, W11261), as described previously(*23*). Briefly, WGA solution (25 μg ml^−1^ in Hanks’ Balanced Salt Solution, HBSS) was allowed to flow through the epithelium channel for 30 min and then washed with continuous flow of HBSS for 30 min. Intestine chips were then cut sideways parallel to the length of the channel and imaged with an epifluorescence microscope (Echo) with ×5 objective.

### Cytokine and chemokine analysis

Apical and/or basal effluents from Intestine Chips were collected and analyzed using a custom panel for quantification of multiple cytokines and chemokines associated with epithelial and endothelial inflammation, including IL-6, IL-8, MCP-1, SDF-1α, PIGF-1, CXCL10, LIF, CCL5, GM-CSF, CXCL-1 IL-22, IFN-γ, TNF-α, MIP-1α and MIP-1β (Procartaplex, Invitrogen). Effluent concentrations were determined using a Luminex 100/200 Flexmap3D instrument coupled with the Luminex XPONENT software.

### NemoCAD predictions

NemoCAD is an AI-based computational tool that can analyze transcriptional changes, that can be targeted to reverse overall transcriptomic changed observed in certain stressors, such as irradiation(*18, 19, 42*). Using this agnostic approach, NemoCAD uses previously defined gene-gene, drug-gene interaction probabilities and differentially expressed gene signatures between control and irradiate states. The algorithm identified potential compound (miconazole in this case), that can revert radiation-induced transcriptional signatures.

### Medium throughput drug screening on ileum organoids

Ileum organoids were recovered from matrigel and fragmented in matrigel at 1:3 split ratio as described above. Fragmented organoids and matrigel were plated in a prechilled U-bottom 96 well plate at 5 µL volume using multichannel p20 pipette. The culture plate was when incubated at 37C for 30 minutes before adding EM+R. 24 hours post plating, culture plates were irradiated. Immediately after radiation, the culture media has been replenished with drugs or DMSO containing the same concentration. Media was changed every 2 days. 7 days post radiation, cell supernatants were collected for cytokine analysis. 100 µL Cell Titer Glo assay 1:1 diluted with PBS (-/-) was added to the plate. Assay has been carried out according to the manufacturer’s protocol. Luminescence was converted to cell counts using a calibration curve created with cells plated at different numbers.

### Transcriptomic Analysis

RNA was extracted from Ileum Chips at 6 hours, 2 days and 7 days after irradiation. Cells in apical and basal channels were harvested separately with RLT lysis buffer from RNeasy Mini Kit (74106; Qiagen, Hilden, Germany), as previously described(*43*). Channels were washed with 100 µL DPBS three times and then fibroblasts were lysed by quickly pressing and releasing the micropipette plunger into the input port of the lower channel at least four times. Lysates were collected in 1.5 mL tubes and stored immediately at −80°C for RNA sequencing analysis. Subsequently, epithelial cell lysates were collected and processed in a similar manner. High-grade RNA (RIN > 8) was prepared for transcriptomic analysis (Agilent Nano Kit 5067-1511 (Agilent 2100 Bioanalyzer, Agilent Technologies) and mRNA sequencing was performed by Azenta Life Sciences (Burlington, MA) via polyA selection using an Illumina HiSeq for 150 bp paired-end reads. Trimmomatic v.0.36 was used to remove possible adapter sequences and nucleotides with poor quality from sequence reads. STAR aligner v.2.5.2b was then used to map the trimmed reads to the *Homo sapiens* GRCh38 with ERCC genes reference genome available on ENSEMBL. Unique gene hit counts were calculated using feature counts from the Subread package v.1.5.2. Only unique reads that fell within exon regions were counted. For plotting of sample expression levels, gene counts were CPM normalized using edgeR v4.2.1(*44*).

Before differential expression analysis, genes were filtered to include only genes with at least 3 reads counted in at least 20% of samples in any group. Differential expression analysis was then performed with the DESeq2 R package(*45*), which tests for differential expression based on a model using the negative binomial distribution, and a sequencing batch was included in the design as a covariate. The false discovery rate (FDR) method was applied for multiple testing correction(*46*). Gene set enrichment analysis (GSEA) was using the fgsea R package and the fgseaMultilevel() function(*47*). The log_2_ fold change from the differential expression comparison was used to rank genes. C5: Gene Ontology gene sets - biological process gene set, C2: Canonical pathways – REACTOME, and C2: Canonical pathways - KEGG collections from the Molecular Signatures Database (MSigDB) (*48, 49*) was curated using the msigdbr R package. Prior to running GSEA, the list of gene sets was filtered to include only gene sets with between 5 and 1000 genes. Differential expression and GSEA were performed using Pluto (https://pluto.bio).

The webapp g:Profiler(*50*) was used to perform functional enrichment analysis of differentially expressed genes. For each comparison, upregulated genes (p-adjusted < 0.05; log2fold change > 1) and downregulated genes (p-adjusted < 0.05; log2fold change < 1) were separately queried using g:Profiler across all data sources. The statistical data scope included only annotated genes and the g:SCS method was used for computing multiple testing corrections for p-values at a threshold of p<0.05. The R package ggplot2 (v3.5.1)(*51*) was used to generate dot plots of gene ontology terms from g:Profiler.

## Statistical Analysis

Data in all graphs are expressed as mean ± standard deviation (SD) and significant differences between multiple groups were determined using an unpaired Students t-test, one-way analysis of variance (ANOVA) or two-way ANOVA with Tukey test for correction unless otherwise indicated in the figure legends. P<0.05 was considered significant. Data were analyzed using GraphPad Prism 8 software (GraphPad Software, San Diego, CA).

## Data Availability

The datasets and analysis will be available upon request.

## Acknowledgement

This project has been funded in whole or in part with Federal funds from the Biomedical Advanced Research and Development Authority (BARDA), Administration for Strategic Preparedness and Response (ASPR), Department of Health and Human Services (DHHS), under Contract No. 75A50123D00004t o D.E.I. and by the Wyss Institute for Biologically Inspired Engineering at Harvard University as well as by an NIH training grants (5T32DK007199-44 and 5T32EB016652-10 to A.Ö.) and Wyss Technology Development Fellowship to A.Ö. Authors would like to thank Yucheng Man and Amanda Jiang for irradiating the samples. We thank Harvard Digestive Diseases Center Organoid Core for providing L-WRN conditioned media (P30DK034854).

## Contributions

A.O. and D.E.I designed the research; A.O., G.E.M, J.P., A.N., N.T. L., T. M., E.C.P., K.H., and A.N., R.R.P. performed the experiments; A.O., G.E.M., J.P., A.N., T. M. analyzed and interpreted the data; A.O., G.E.M. and J.P. established ileum human organoid biobanks; M. S., R. G. and A.O. performed bioinformatic analysis; D.B.C., R.R., L.B. and D.B. provided the clinical samples; A.O. and D.E.I. acquired the funding; A.O. wrote the articled with input from G.G., and D.E.I.; and all authors reviewed, discussed and edited the manuscript.

## Data and code availability

The datasets and analysis will be available upon request.

---

**Extended Data Figure 1:**
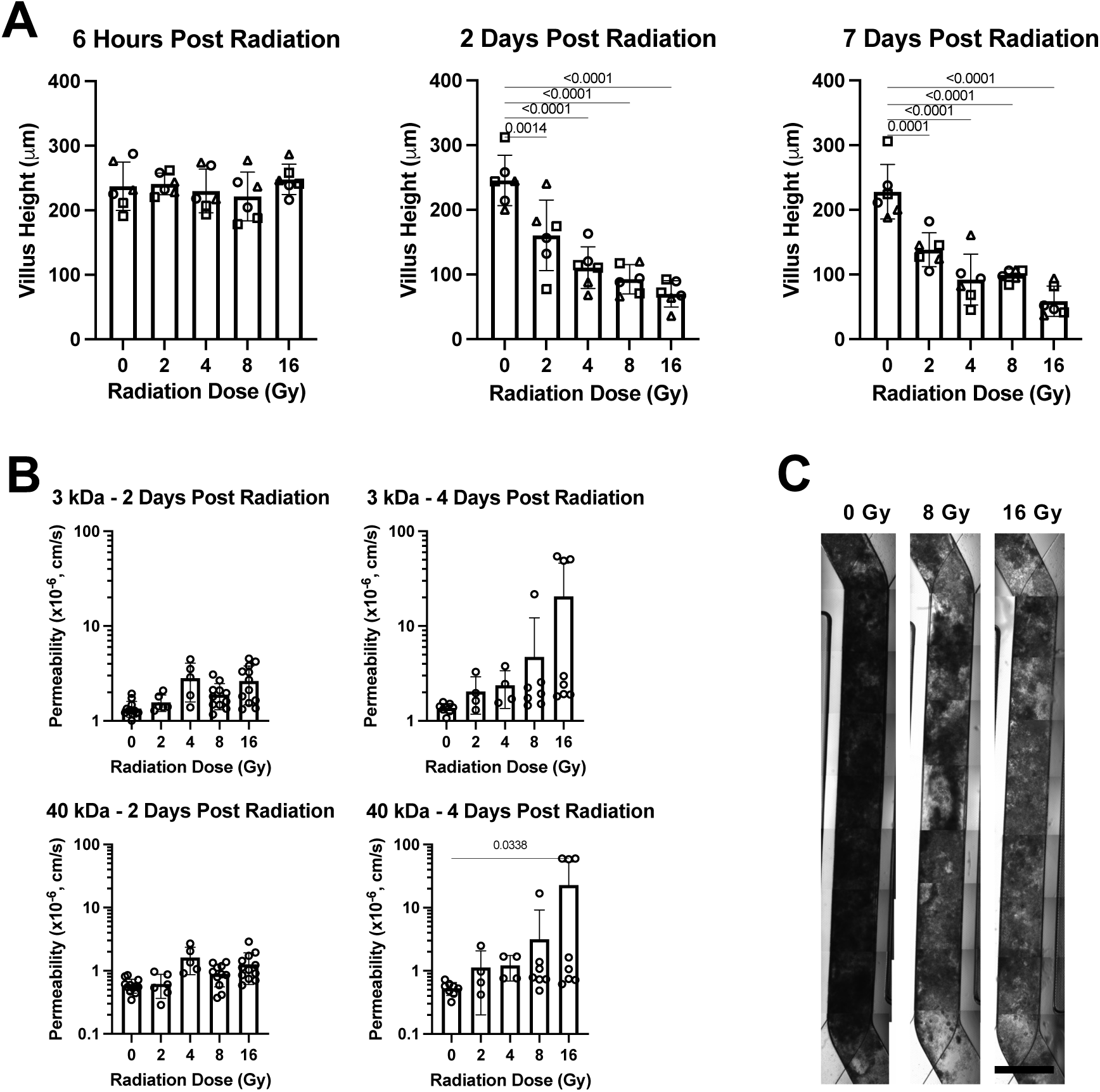
Radiation dose dependent villus blunting and tissue barrier leakiness of Ileum Chips. **A)** Villus height in Ileum Chips exposed to different radiation doses. Villus blunting begins 2 days post-radiation and increases by day 7 after irradiation (n = 3 healthy donors).**B)** Graphs showing that intestinal barrier permeability to Cascade Blue is significantly higher in Ileum Chips exposed to 16 Gy radiation. The increase in permeability to 40 kDa tracer is observed on day 4 (n = 3 donors). **C)** Representative transmitted light images of whole Ileum Chips representative of 3 donors 7 days after being exposed to 0, 8 and 16 Gy radiation doses. A functional epithelial monolayer with accumulated mucus, which appears as opaque blackened fuzzy material, in the apical channel of all chips over the 7 day; however, accumulated mucus was partially lost in different regions of Ileum Chips were exposed to 8 or 16 Gy radiation (Bar, 1 mm). Each symbol represents an individual health patient derived chip.

**Extended Data Figure 2:**
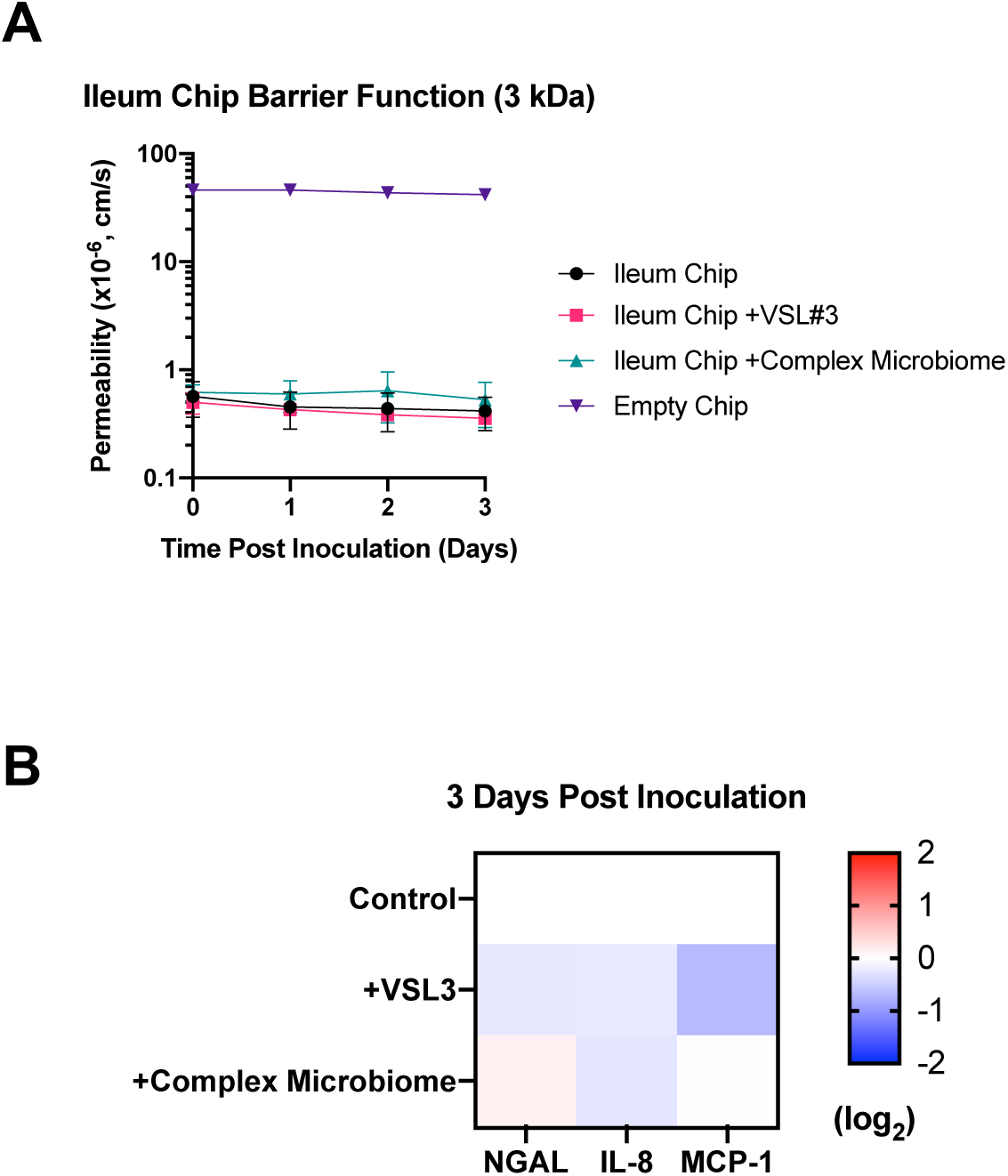
Culturing VSL#3 or complex microbiome does not interrupt tissue barrier integrity of Ileum Chips. **A)** Changes in barrier integrity measured by quantifying Cascade blue transport across the tissue−tissue interface within the Ileum Chip cocultured with complex microbiome or VSL#3 probiotic consortium under anaerobic conditions (n = 4 individual chips; data are presented as mean ± s.d.). Co-culture with different microbiome species did not interfere the barrier integrity. **B)** Inflammatory cytokine production assessed using the outflows collected from the apical channel. Co-culture with different microbiome did not induce inflammatory cytokine production (n = 4 individual chips).

**Extended Figure 3:**
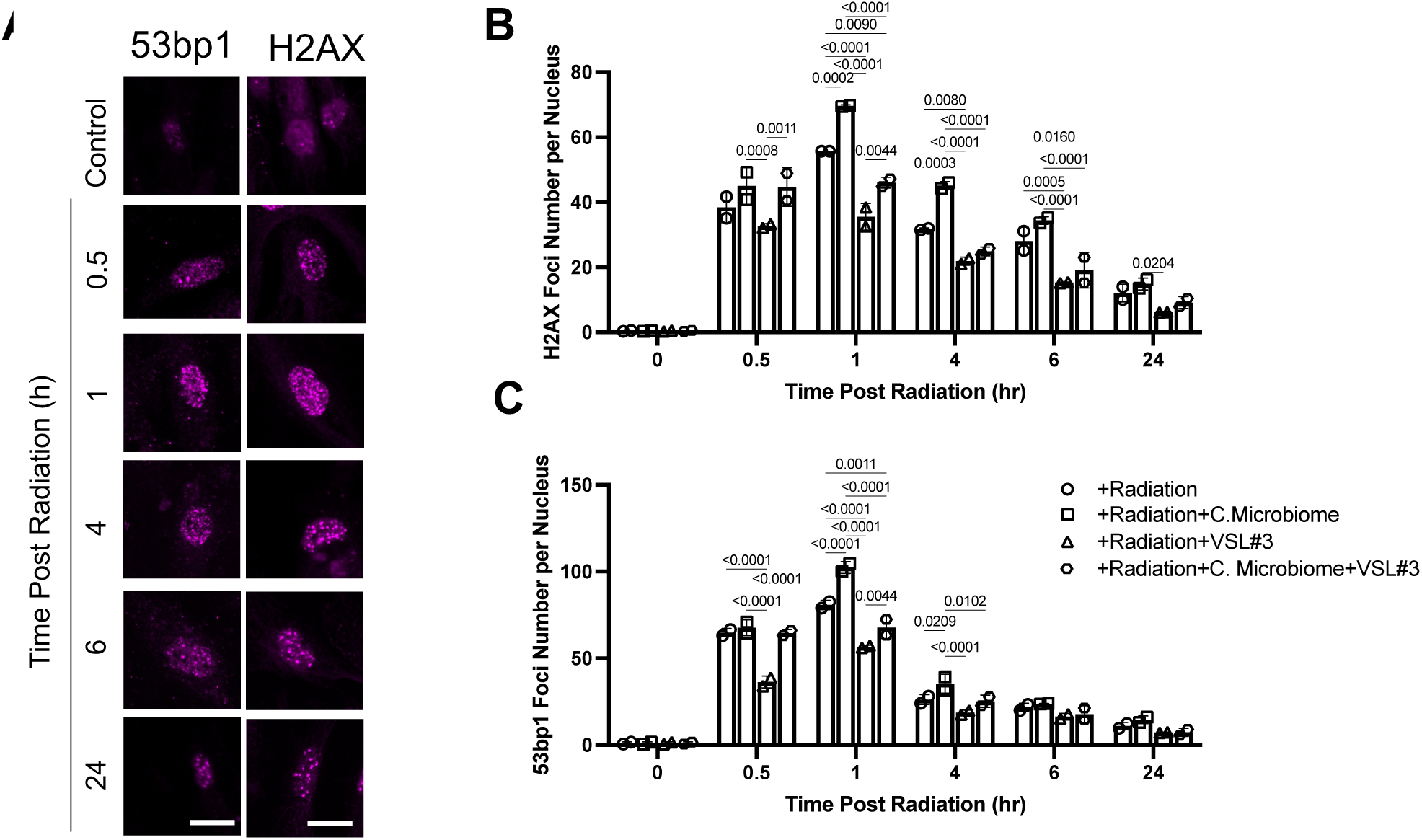
Time-course analysis of DNA fragmentation in endothelial cells cultured in Ileum Chips with complex microbiome and the VSL#3 probiotic consortium following γ radiation exposure. **A)** Time course analysis of radiation effects on formation of DNA double-strain breaks as detected by increased punctate staining of H2AX (indicative of DNA damage) and 53bp1 (indicative of impairment of DNA damage repair) in endothelial cell cultured in Ileum Chips. Nuclear foci formation of both markers maximizes 1hr post radiation and decreases over time (Bars are 10 µm). **B)** Quantification of radiation-induced time-course changes in H2AX and **C)** 53bp1 foci number per nucleus in the presence of complex microbiome, VSL#3 or both. Complex microbiome increases radiation-induced H2AX and 53bp1 foci number in nucleus. VSL#3 probiotic consortium prevents complex microbiome induced nuclear H2AX foci formation in the first 6 hours. VSL#3 prevents nuclear 53bp1 foci formation in the first 4 hours. Numbers indicate P values between compared groups, as determined by one-way ANOVA test (n=2 Healthy donors from 2 independent experiments). Each symbol represents an individual health patient derived chip.

**Extended Data Figure 4:**
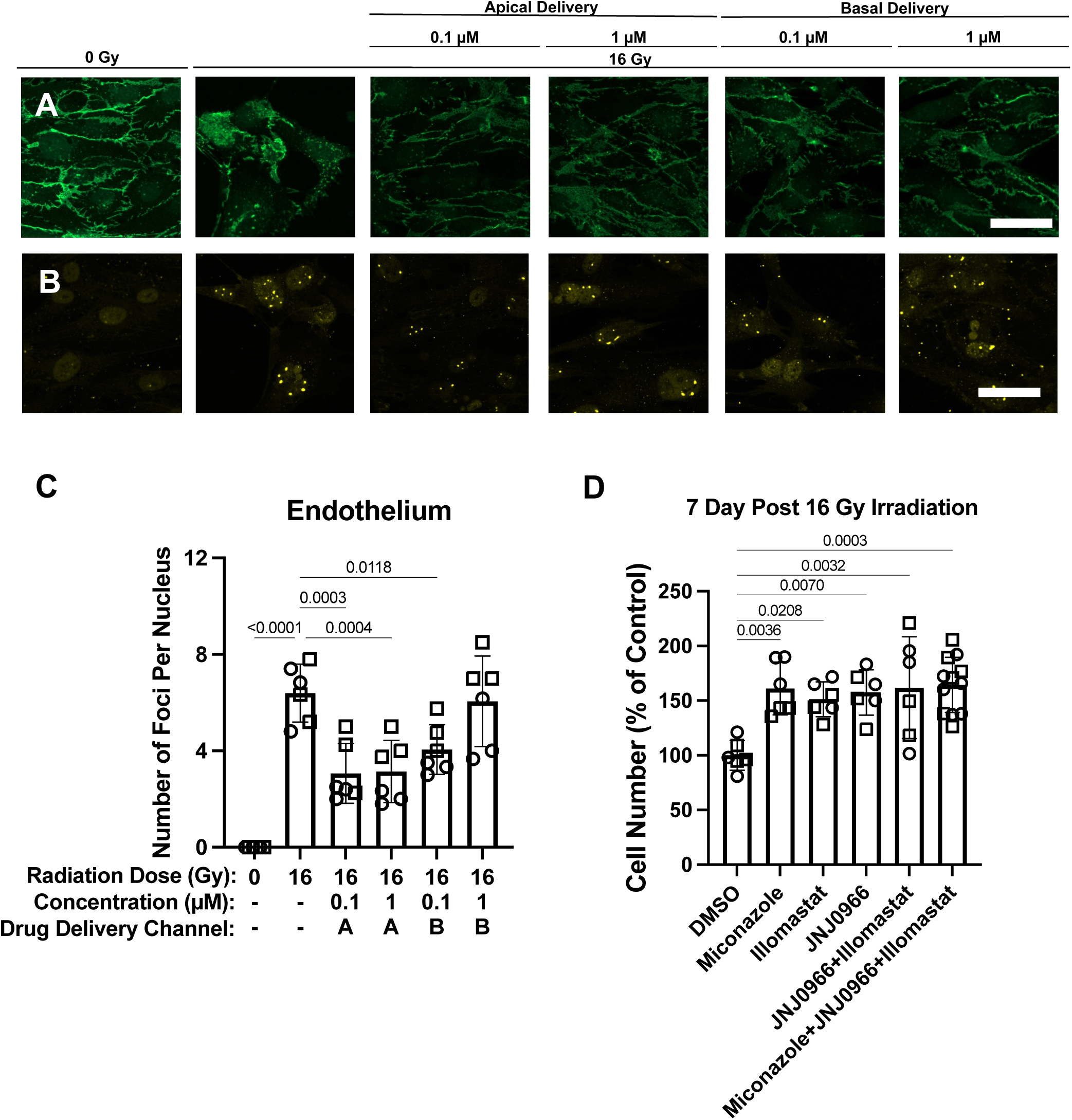
Miconazole’s radioprotective mechanism may Involve MMP9 inhibition. **A)** Immunostaining with VE-cadherin (endothelial cells, green), showed that junction disruption due to 16 Gy irradiation could be partially prevented with miconazole treatment (Bar, 20 μm). **B)** Quantification of nuclear 53bp1 foci (yellow) in endothelial cells cultured in Ileum Chips and treated with miconazole after 16 Gy irradiation. **C)** Quantification of foci count per nucleus presented in B. **D)** Quantification of cell numbers in 16 Gy irradiated ileum organoids and treated with different inhibitors suggests that miconazole’s radioprotective effect may be mediated through MMP9 inhibition (n = 2 donors). Each symbol represents an individual health patient derived chip.

